# Community engaged tick surveillance and *tickMAP* as a public health tool to track the emergence of ticks and tick-borne diseases in New York

**DOI:** 10.1101/2022.02.17.22271128

**Authors:** Charles E Hart, Jahnavi Reddy Bhaskar, Erin Reynolds, Meghan Hermance, Martin Earl, Matthew Mahoney, Ana Martinez, Ivona Petzlova, Allen T Esterly, Saravanan Thangamani

**Author notes:** These first authors contributed equally to this article (Charles E Hart, Jahnavi Reddy Bhaskar, Erin Reynolds).

## Abstract

A community engaged passive surveillance program was utilized to acquire ticks and associated information throughout New York state. Ticks were speciated and screened for several tick-borne pathogens. Of these ticks, only *I. scapularis* was commonly infected with pathogens of human relevance, including *B. burgdorferi*, *B. miyamotoi*, *A. phagocytophilum*, *B. microti*, and Powassan virus. In addition, the geographic and temporal distribution of tick species and pathogens was determined. This enabled the construction of a powerful visual analytical mapping tool, *tick*MAP to track the emergence of ticks and tick-borne pathogens in real-time. The public can use this tool to identify hot-spots of disease emergence, clinicians for supportive evidence during differential diagnosis, and researchers to better understand factors influencing the emergence of ticks and tick-borne diseases in New York. Overall, we have created a community-engaged tick surveillance program and an interactive visual analytical *tick*MAP that other regions could emulate to provide real-time tracking and an early warning for the emergence of tick-borne diseases.

## INTRODUCTION

Tick-borne pathogens continue to emerge and are a major health concern in the United States. The most common of these include *Borrelia burgdorferi*, *Anaplasma phagocytophilum*, and *Babesia microti*, the causative agents of Lyme disease, human granulocytic anaplasmosis (HGA), and Babesiosis respectively. The existence of these pathogens has been known for over forty years, continued tick surveillance has identified new and emergent tick-borne pathogens over the last two decades. These include *Borrelia miyamotoi* (1), *Ehrlichia muris eauclairensis* (2), *B. mayonii* (3), Heartland virus (4), Bourbon virus (5) and Powassan virus (POWV) (6). In comparison to well characterized pathogens such as *B. burgdorferi*, newly discovered pathogens are less understood in regard to prevalence and human disease burden. The frequency with which new tick-borne pathogens have been discovered suggests that additional pathogens may also exist but remain undiscovered. Interactions between these pathogens and changes in their range and sylvatic cycle may also be actively occurring, compounding the complexity of tick-borne disease in New York State.

Tick surveillance is useful for assessing the geographic and temporal distribution of tick-borne pathogens and vector species, as well as for identifying patterns that can be used to predict changes that may affect the transmission of important pathogens. In active surveillance, a specific area is flagged for ticks to assess tick density, species presence, and pathogen prevalence rates in association with a specific environment. While this method provides a more complete understanding of a known ecosystem, it is limited in scale to smaller areas requiring extensive manpower to study. The alternative is passive surveillance, where ticks are submitted by individuals living in a specific region rather than having researchers collect them. This allows for access to ticks from much wider geographic areas with lower collection manpower. For the assessment of rare pathogens, both systems may be combined, using passive surveillance to identify geographical foci where the pathogen is present to choose locations for more intensive active collection. Although potential biases and limitations must be taken into account with passive surveillance, these systems provide an invaluable tool for understanding where and when contact with ticks, and by extension tick-borne pathogens, occurs. In this work, we describe community engaged tick surveillance and the development of an interactive mapping tool, *tick*MAP, to track the emergence of ticks and tick-borne disease-causing agents in New York.

## METHODS

### Tick submission and data collection

The ticks utilized for this analysis includes those submitted between April and December of 2020, with the online-available mapping tool including ticks up to the present. Advertising of our tick surveillance was accomplished through the laboratory website, local news coverage, pamphlets handed out at the New York State Fair, social media, and subsequently propagated by word-of-mouth.

Tick receipt was documented using a two-part online survey in REDCap (7,8). Part one of the survey, the Tick Submission Form, was completed by public tick submitters. This questionnaire was approved by the SUNY Upstate Medical University Institutional Review Board (Protocol 1739668-1) This form included a series of questions to identify the geographic location and when tick exposure occurred in addition to the host species and tick feeding site (Supplementary Figure 1A). A unique tick ID number was generated for each tick when the tick submitter completed the tick-submission form.

The survey form records the location of the tick as entered, which is relayed by the submitter. As there is a possibility that the tick may have been acquired in a secondary location and inadvertently transferred either on an individual, a pet, or property, before being discovered and submitted, the survey form provides instructions to the submitter to provide us the location where the human or pet may have encountered the tick.

Part two of the survey, the Lab Form (Supplementary Figure 1B), was completed by laboratory personnel upon receipt of a specimen and had prepopulated fields and drop-down menus to streamline the receipt and identification process. Pathogen results were entered into REDCap and verified before releasing to the tick submitters. Throughout this process, automated emails are sent to the email address provided by the submitter when each of the following activities was completed: 1) The Tick Submission Form was completed by the tick submitter, 2) The tick sample was received in the lab and the Lab Form populated for the tick sample, and 3) Pathogen results were verified.

### Tick identification

Ticks were morphologically identified, surface decontaminated with 70% ethanol followed by rinsing with deionized water (9–14). Samples which could not reliably be identified due to damage, decomposition, or morphology inconsistent with published information were identified to genus level or designated as unknown when genus could not be determined. Fed-status was recorded if observable or if the submission form had identified the tick as having been found attached to a host. Samples were stored at approximately 4°C in Schneider’s *Drosophila* Medium (Gibco, Paisley, UK) supplemented with 9% fetal bovine serum (Hyclone, Logan, UT.)

### RNA Extraction

This study utilized an RNA-based protocol for the detection of pathogens. This allowed for simultaneous detection of eukaryotic, prokaryotic, and viral pathogens with identical processing steps for each tick. Individual ticks in tubes containing 300 to 500μL media were homogenized using a Qiagen Tissue Lyser II operating at 30 cycles/second for five minutes. Ticks that visually did not homogenize completely received a second identical round of homogenization. The homogenate was briefly centrifuged to remove debris before 150µL of the supernatant was removed and mixed with 200μL RLT buffer (Qiagen, Germantown, MD). The remaining homogenate was stored at approximately -80°C to provide the option of culturing pathogens in future studies. The RNA extraction process was performed using a QIAcube HT (Qiagen, Germantown, MD) automatic extraction robot and Qiagen RNeasy 96 QIAcube HT kits according to the manufacturer’s instructions.

### Detection of pathogens by qRT-PCR

For detection of pathogen RNA by rt-qPCR, a Reliance One-step Multiplex Supermix (Bio-Rad, Hercules, CA) was used, with 5μL of kit master mix combined with 4.5μL of either primer combination “A” or “B”, using the sequences listed in Table 1. Ten and a half microliters of extracted tick RNA was used for each test, loaded mechanically from the QIAcube output plates with a Qiagen QIAgility robotic plate loader. Control DNA fragments were synthesized by (Azenta, formerly Genewiz, South Plainfield, NJ, USA) to include the area of the target sequence enclosed by the primers listed, along with an additional flanking region of 20bp on either end. Equal quantities of each of the appropriate primer-specific sequence tested were combined into controls for primer mixtures mixes “A” and “B”. Each plate was run with one positive control and one no template control (NTC). PCR was run using CFX96 Touch Real-time qPCR systems (Bio-Rad, Hercules, CA). The cycle began with 50°C for 10min, followed by 95°C for 10min. Then 45 cycles of 95°C (10sec) and 60°C (30sec) were conducted, with fluorescence measured in the 60°C step using the all the excitation/emission frequencies allowable by the machine’s internal color filters, optimized for FAM, HEX, TEX-615 (Texas Red), CY-5, and CY-5.5.

**Table 1.**
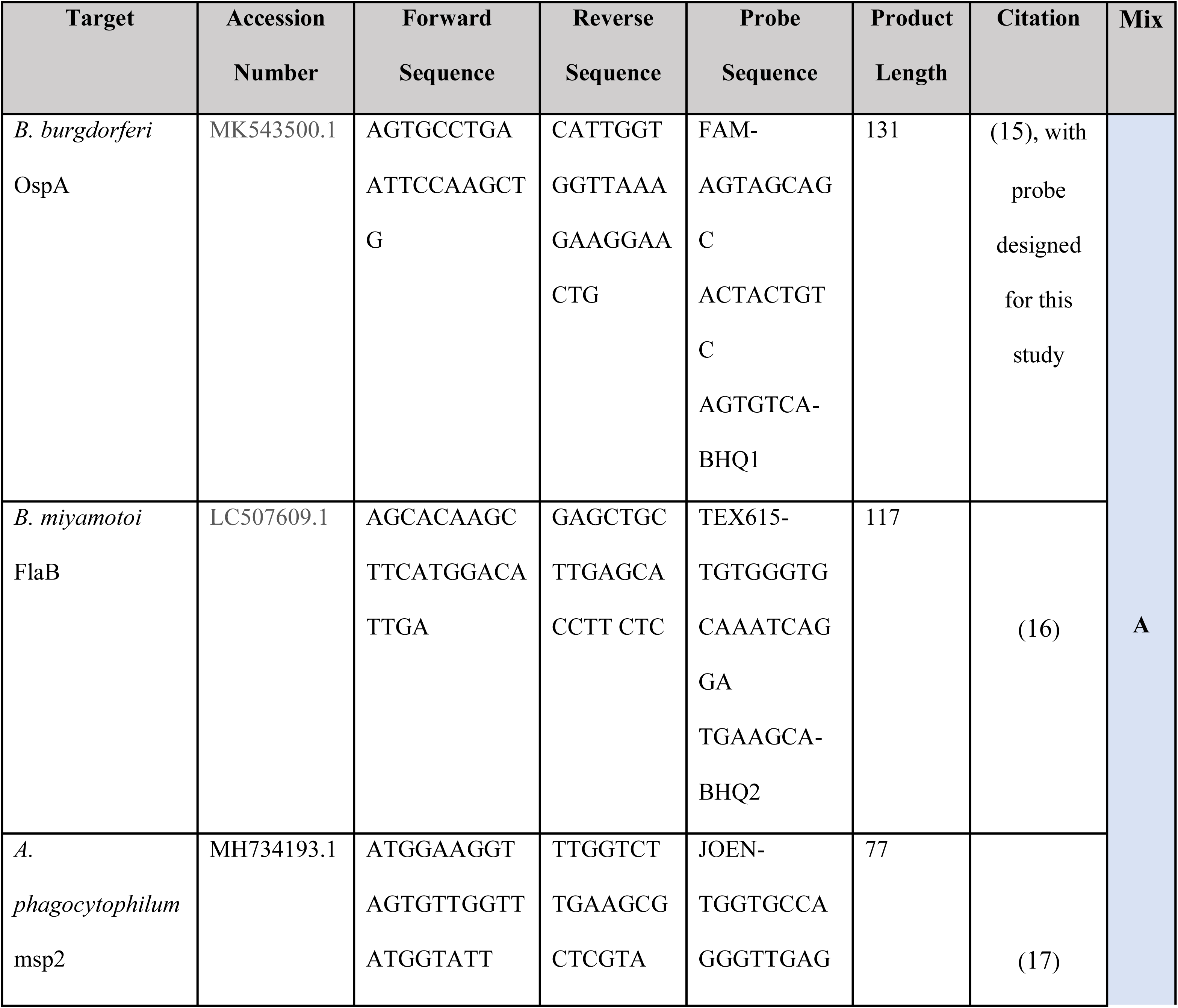
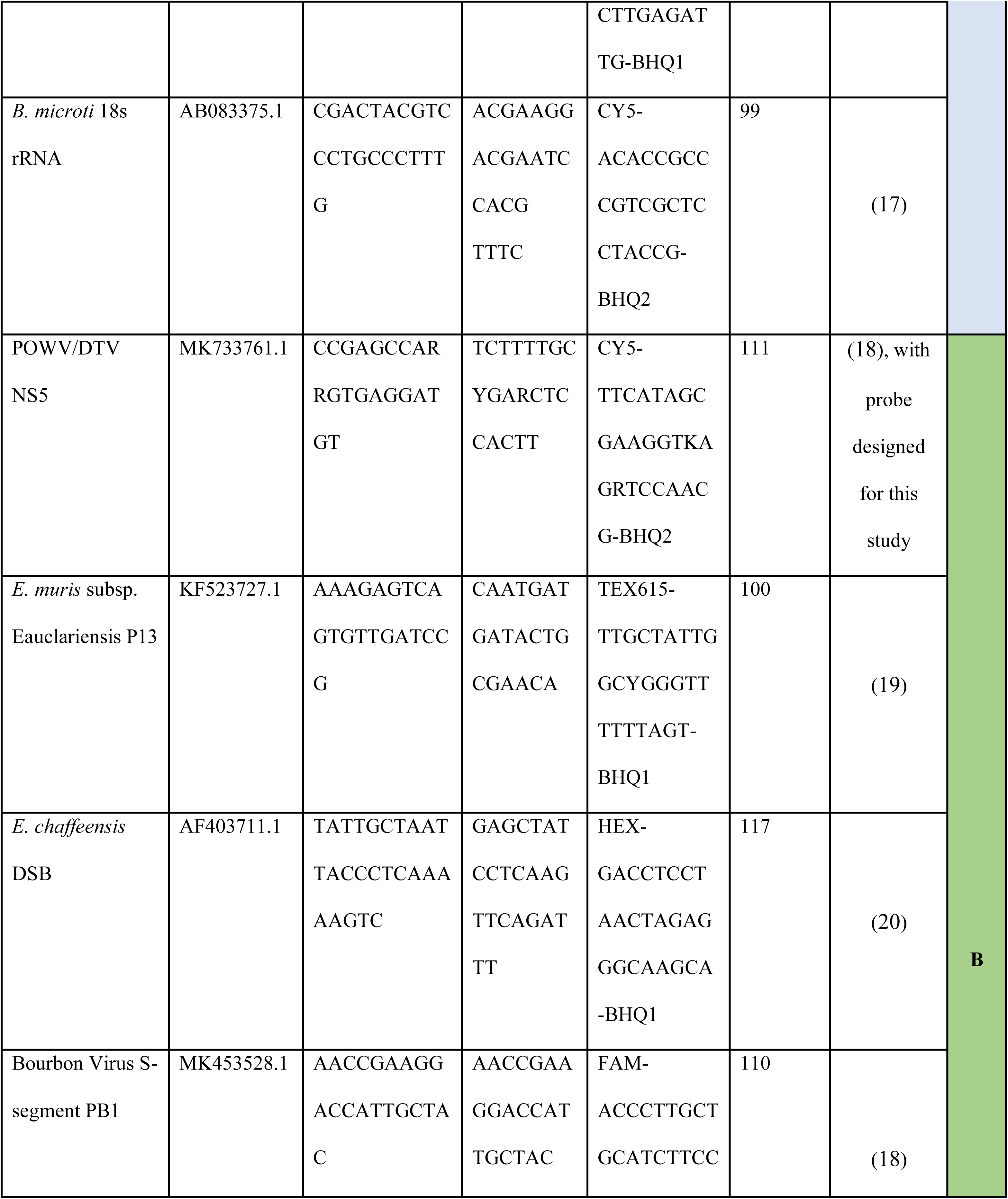
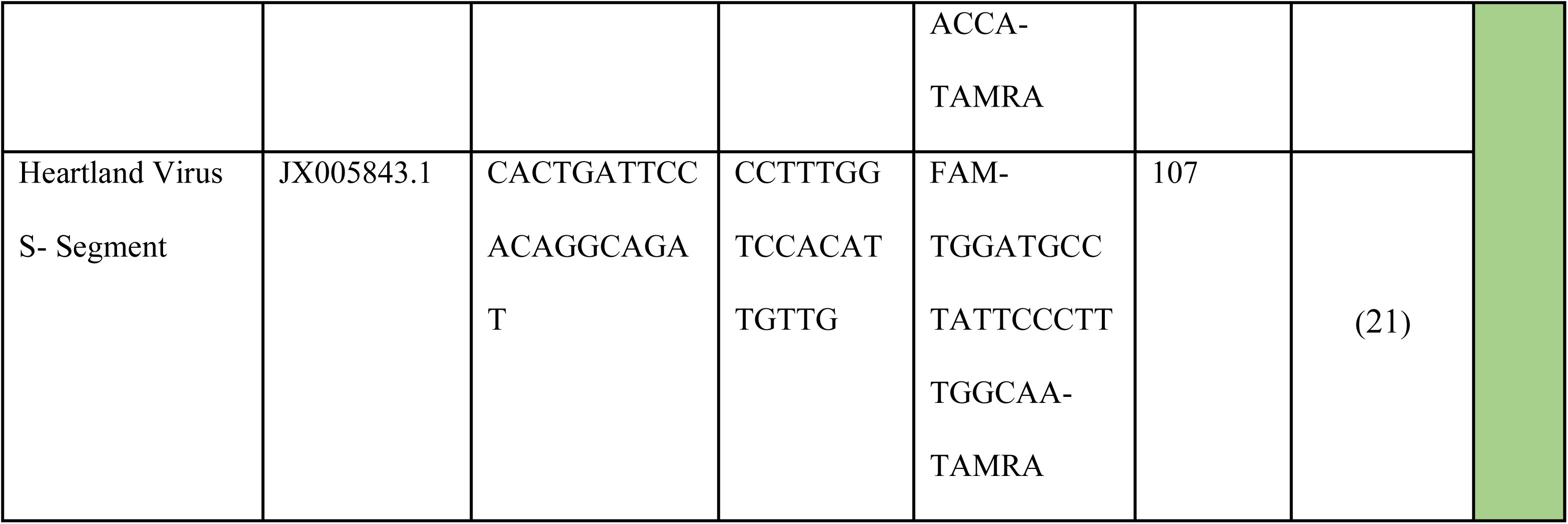
Primers Used for Pathogen Detection. The target genes, primer sequences, probe sequences with fluorophore/quencher combinations, and product lengths of the pathogen-detection primers used in this study. These were combined in to Mix A and Mix B, with each tick sample run once with each primer combination to assess the presence of nine pathogens.

### Interactive mapping tool dashboard (tickMAP )

Mapping tools to track the distribution of ticks and tick-borne pathogens have been helpful in gaining relevant information directly to understand the abundance or geographical distribution and expansion of ticks and tick-borne pathogens. Here we have utilized professional version of the Tableau Software to generate meaningful visualization for comparison of various tick surveillance data through our interface, *tick*MAP(Mapping Arthropods and Pathogens). Tableau Software uses the data stored in REDcap to create real time dashboards. A workflow was developed for Tableau to communicate with the REDCap database where the tick submission form and lab form are collected. The REDCap database was customized based on the unique workflow and specific data points that needed to be validated by the system to ensure accuracy. As the data is collected, a connection between the REDCap database and Tableau was created to refresh the dashboard daily. Customization was done to create informative and accurate dashboards for quick and easy use. The map displays a variety of information which can be filtered by any specific category. This includes tick species, pathogen species, tick life stage, and the host species on which the tick was found. This user-friendly dashboard is available at www.nyticks.org and www.tickmap.org for the public to view and is updated daily. An example of this interface is displayed in Figure 1. The publicly available map can provide data down to a county level, although the underlying data and Tableau mapping software is capable of rendering the data to the zip-code level. The base map for the images was obtained from Mapbox which is an open source map available at http://www.studio.mapbox.com

**Figure 1.**
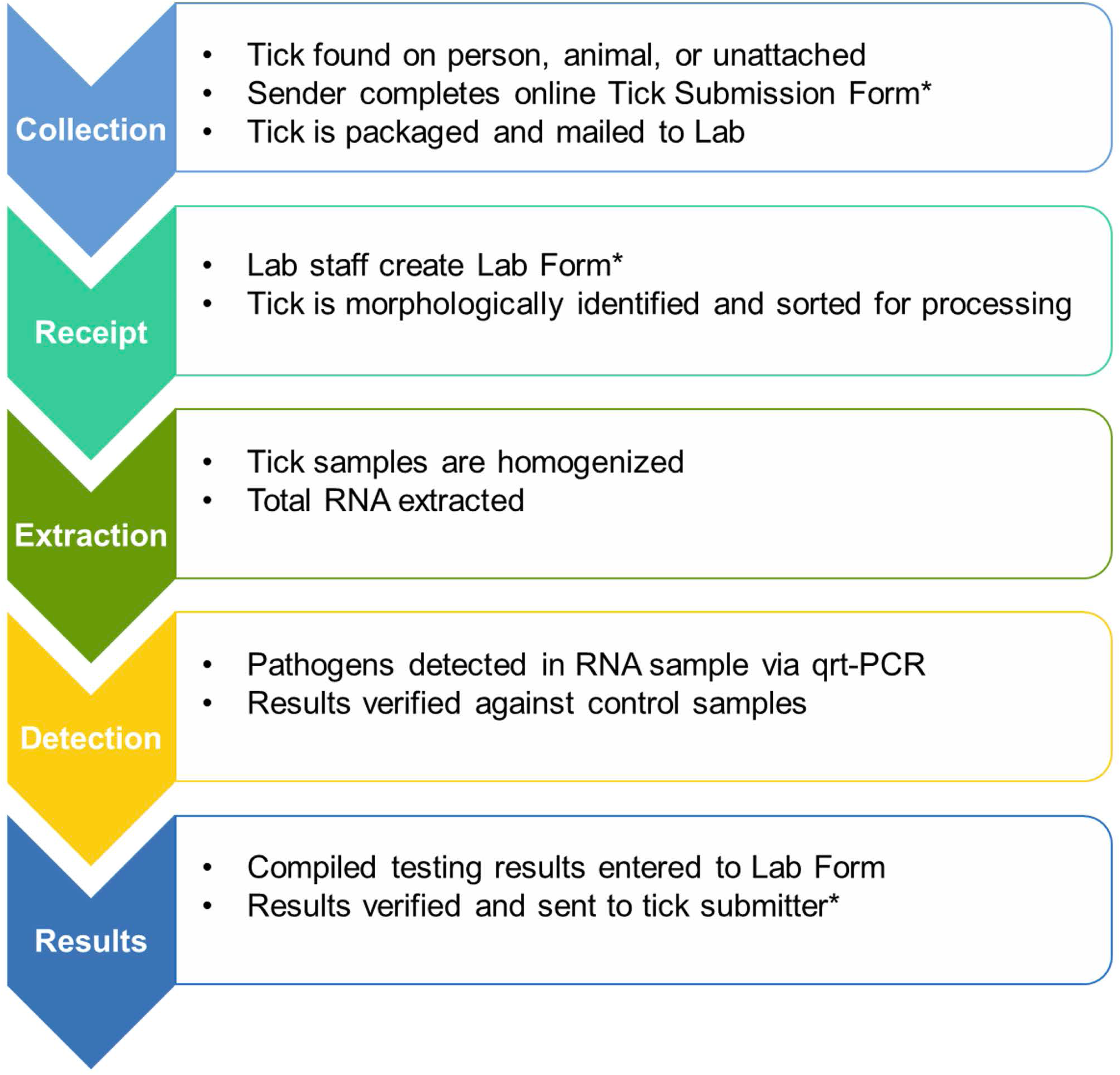
Example of the Public-Facing Web Application. The web-application automatically generates color-coded maps and bar graphs in real-time through a Tableau interface with the REDCap database containing tick information. This application displays tick life stages, species, and pathogen and can be filtered by location, time period, tick species, tick lifestage, or by pathogen.

An overview of the complete workflow of the Citizen Science Program is displayed in Figure 2.

**Figure 2.**
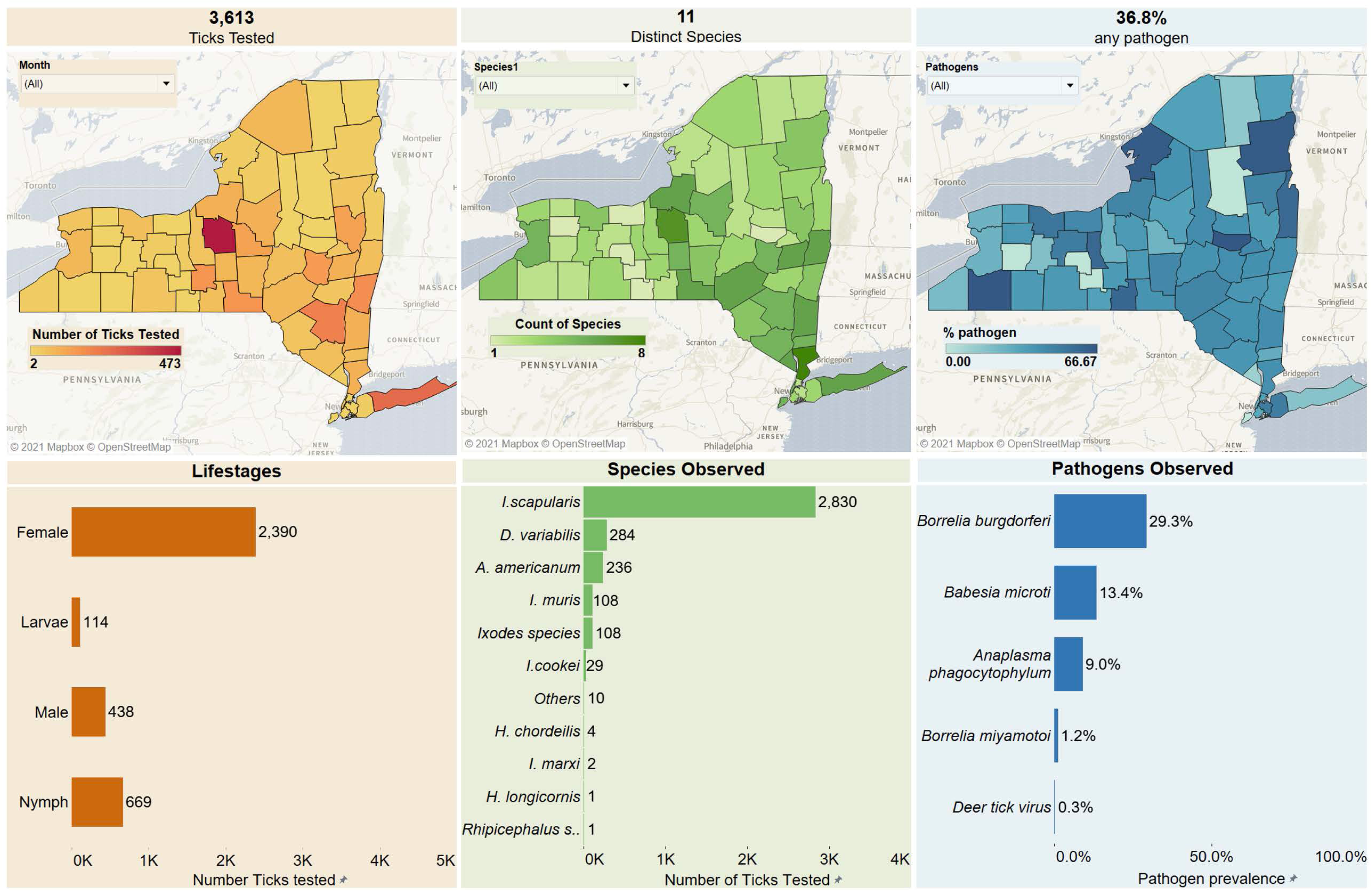
Overview of Tick Surveillance Workflow. Tick surveillance sample processing covers five major functions, each with multiple steps. At steps designated with (*), an automatic notification is sent to the email linked to the Tick ID to provide status updates to the tick submitter.

## RESULTS AND DISCUSSION

### Tick Submissions were Dominated by *Ixodes*, *Dermacentor*, and *Amblyomma*

A total of 3613 ticks were received between April and December 2020 (Figure 3). Of these, 2830 (78%) were *Ixodes scapularis*, 284 were *Dermacentor variabilis* (8%) and 236 were *Amblyomma americanum* (7%). An additional 249 represented other *Ixodes* species, including 108 *I. muris*, 29 *I. cookei*, two *I. marxi*, and 108 *Ixodes* spp. ticks which could not be identified to species level. One *Rhipicephalus sanguineu*s tick and five *Haemaphysalis* ticks were received. Of the *Haemaphysalis* ticks, four were identified as *H. chordeilis*, a native species and one was *H. longicornis*, a recently-introduced invasive species native to Asia.

**Figure 3.**
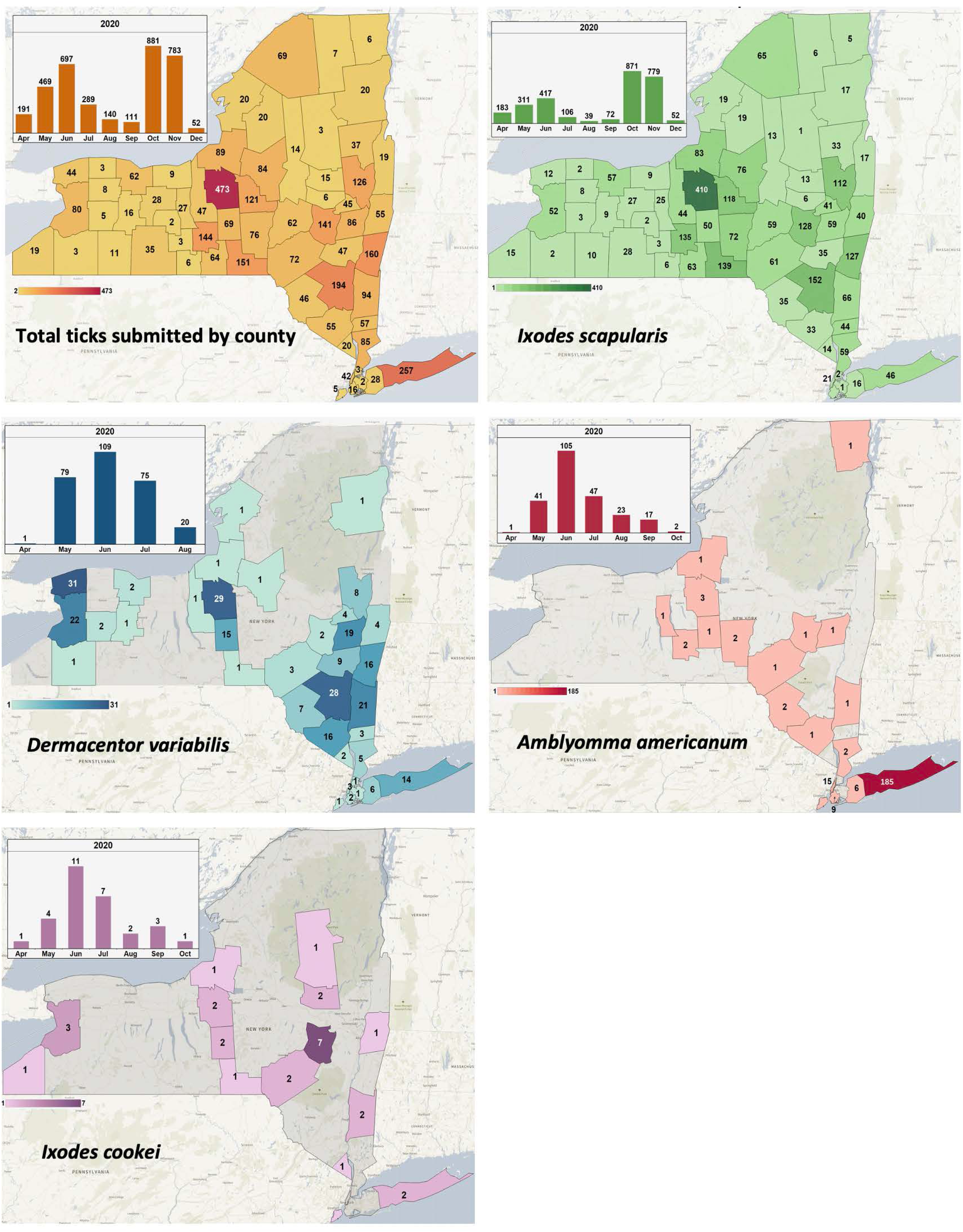
Distribution by County of the Total Ticks Tested Through the Citizen Science Program. The spatial and temporal distribution of ticks indicates that *I. scapularis* is distributed throughout New York, with peak submissions during the spring and fall. *D. variabilis*, *A. americanum*, and *I. cookei* were submitted primarily during the summer and most often from the southern portion of the state, with *D. variabilis* additionally having a high submission rate in the west and *A. americanum* having its highest concentration in Suffolk County, Long Island.

The division of lifestages received was dependent on species. *Ixodes scapularis* was received as adults, nymphs, and a small percentage of larvae, with adult females being the most common lifestage. *Amblyomma americanum* was received primarily as nymphs and adults, with nymphs being the most common stage received. *D. variabilis*, in contrast, was exclusively submitted in its adult form supporting that the larval and nymphal stages are not often encountered biting humans or domestic animals. These immature ticks are believed to feed on small rodents such as *Peromyscus* spp. (22) and can be found in association with larval *I. scapularis*, which share a similar host preference (23).

### Host Origin of Ticks Indicates Association with Humans and Domestic Animals

A portion of the submission survey completed by tick submitters included information concerning the host on which the tick was found. These numbers of ticks of known and unknown species submitted from known hosts are presented in (Table 2). The majority of ticks (2193, 60.7%) were identified as having been found on humans. Of these, the majority were *I. scapularis* (1592, 56.92%). A high percentage of *D. variabilis* (159, 55.8%) and *A. americanum* (203, 86.01%) were also submitted from humans, as well as for *I. cookei* despite the low submission (17, 58.6%) submitted.

**Table 2.** Animal Origins of Submitted Ticks. The majority of ticks submitted originated from humans and closely associated animals (pets), with small numbers submitted from horses, deer, and a goat. Some detected ticks were unattached, although most were found on a mammalian host.

| Species | Host fed upon |  |  |  |  |  |  |  | Total |
| --- | --- | --- | --- | --- | --- | --- | --- | --- | --- |
|  | Unattached | Cat | Dog | Deer | Goat | Horse | Human | Unknown |  |
| *I. scapularis* | 436 (15.4%) | 103 (3.64%) | 606 (21.41%) | 12 (0.42%) | 1 (0.04%) | 41 (1.45%) | 1592 (56.25%) | 39 (1.4%) | 2830 |
| *D. variabilis* | 57 (20%) | 3 (1.05%) | 49 (17.2%) |  |  | 16 (5.6%) | 159 (55.8%) | - | 284 |
| *A. americanum* | 6 (2.55%) | - | 23 (9.8%) | 1 (0.45) | - | 1 (0.42%) | 203 (86.01%) | 2 (0.84%) | 236 |
| *H. chordeilis* | - | - | - | - | - | - | 4 (100%) | - | 4 |
| *H. longicornis* | - | - | 1 (100%) | - | - | - | - | - | 1 |
| *I. marxi* | - | - | - | - | - | - | 2 (100%) | - | 2 |
| *I. muris* | 5 (4.6%) | 1 (0.9%) | - | - | - | - | 102 (94%) | - | 108 |
| *I. cookei* | - | 5 (17.2%) | 7 (24.1%) | - | - | - | 17 (58.6%) | - | 29 |
| *Ixodes* species | 3 (2.8%) | - | 1 (0.9%) | - | - | - | 104 (96.3%) | - | 108 |
| *Rhipicephalus* spp. | - | - | 1 (100%) | - | - | - | - | - | 1 |
| Others | - | - | - | - | - | - | 10 (100%) | - | 10 |
| Total |  |  |  |  |  |  |  |  | 3613 |

The next two most common hosts from which ticks were submitted were dogs (688, 19.5%) and cats (112, 3.1%). In both cases, *I. scapularis* was the primary species of ticks submitted, with 606 (21.41%) *I. scapularis* submitted from dogs and 103 (3.64%) submitted from cats. For submitted *D. variabilis*, 49 (17.2%) were submitted from dogs while only 3 (1.05%) were from cats. A small portion of the submitted *A. americanum* were acquired from dogs (23, 9.8%), and none were from cats.

A relatively low number of ticks were submitted from horses 58 (1.6%). These consisted primarily of 41 *I. scapularis*, although making up only 1.45% of the total *I. scapularis* submitted. Sixteen *D. variabilis* were submitted, constituting 5.6% of all *D. variabilis* submitted. Only one *A. americanum* was submitted from a horse. Thirteen ticks were also submitted from deer, with twelve of those being *I. scapularis* and one being *A. americanum*. One *I. scapularis* was submitted from a goat. Ticks were either not submitted from other domestic or wild animals or host information was not provided.

This pattern of identified tick hosts is primarily influenced by the manner that the public encounters ticks rather than the host specificity of each species. Humans are likely more concerned with ticks found on themselves and pathogens and the possibility of these ticks transmitting human-infecting pathogens, hence the bias of the submissions toward ticks submitted from humans. Submissions from animals require that the animal have contact with humans to detect and submit the tick, and also is restricted to animals that have access to tick habitats. These are primarily domestic animals, including pets (dogs and cats) and farm animals with close human contact (horses). Samples from wild animals such as deer and lower human-contact farm animals such as goats were seldom submitted. While the survey does not include a response option for how the deer were encountered, hunting or animal rescue are the most likely sources.

The most common species of tick from any host was *I. scapularis*, with *D. variabilis* and *A. americanum* as the second and third most common. This reflects the tick submission data present throughout the state from any source. *D. variabilis* was more common on dogs than cats, and made up a high proportion of the ticks present on horses, although the limited number of ticks submitted from cats and horses makes the actual rate of the less-common tick species difficult to assess. *A. americanum* was identified primarily from humans and dogs, with one submitted from a deer and one submitted from a horse. This suggests that these ticks are less commonly encountered on pets than other tick species and are more anthropophilic than other species. This may relate to either the actual preferences of the species or the dynamics of human-tick interactions specific to highly-populated Long Island where *A. americanum* is most common.

These data indicate that while human encounters with ticks are common, domestic, peri-domestic and wild animals also encounter pathogen-transmitting species of ticks. These animals are susceptible to infection by tick-borne agents tested in this study. Dogs and horses can exhibit clinical manifestations of Lyme disease when infected with *B. burgdorferi*, although cats usually experience subclinical illness (24,25). Equine and canine anaplasmosis can also occur in response to infection with *A. phagocytophilum* (26,27), although as with *B. burgdorferi* cats, are less likely to develop acute illness when infected (28). The role of *B. microti* specifically is poorly understood with regard to veterinary significance. Other *Babesia* species are responsible for symptomatic infection in cats and horses rather than *B. microti*. These include *B. felis* and *B. cati*, among others, none of which are considered important veterinary pathogens in North America (29). Equine babesiosis is generally due to infection with *B. caballi* or *Theileria cervi* (30). Canine babesiosis, likewise, is usually caused by *B. canis* (31), although *microti*-like *Babesia* have been detected in dogs in Europe and can produce severe, life-threatening infection (32). The veterinary significance of *B. miyamotoi* has not been widely assessed, although it has been detected in the blood of healthy cats (33), suggesting that cats are capable of experiencing subclinical infection.

Of particular veterinary importance for emergence is the piroplasm *Cytauxzoon felis*, a parasite transmitted by *A. americanum* in the southern half of the United States (34). This disease causes organ failure and death in felines and exists in sylvatic cycles using the bobcat (*Lynx rufus*) as a reservoir. While the disease has not been diagnosed in New York, the presence of mainland populations of *A. americanum* suggest the possibility of its establishment in the future. Long Island, where most *A. americanum* were submitted from, is unlikely to be capable of sustaining the sylvatic cycle of *C. felis* due to the expatriation of *L. rufus* from the island.

### Tick Seasonality is Dependent on Species

The number of ticks submitted varied throughout the year (Figure 3). In early spring (April), 191 ticks were submitted, including 183 *I. scapulariş* with only one of each *D*. *variabilis*, *I. cookei*., and *A. americanum*. By May, the number of later-season ticks (*A. americanum* and *D. variabilis*) began to increase; out of 469 ticks submitted, 311 were *I. scapularis* with an additional 79 *D. variabilis*, 41 *A. americanum*, and 4 *I. cookei*. This April-May period corresponds to the re-activation of overwintering ticks. *Ixodes scapularis*, being the more cold-tolerant of these species, emerges early in spring with intermittent activity when temperatures are above freezing. *D. variabilis* and *A. americanum* are less cold-tolerant and did not become fully active until later spring when temperatures were consistently higher.

*Dermacentor variabilis* and *A. americanum* were most commonly submitted during the summer, starting from a peak in June and decreasing to zero by August for *D. variabilis* and October for *A. americanum*. This is partially similar to the pattern observed for *I. scapularis*, which experienced a similar peak in June followed by decreasing submissions until September. Submissions of *I. scapularis* vary from other species in that they exhibit a sudden and substantial surge in October and November. In June, 417 *I. scapularis* were submitted, decreasing to 106 in July, 39 in August, and 72 in September. The submission of adult ticks decreased while the number of nymphs increased, suggesting that egg laying, hatching, and larval feeding likely occurred earlier in the spring and were not observed. These larvae were not observed, as *I. scapularis* larvae prefer feeding on small rodents such as *Peromyscus* spp. (23) and are less likely to interact with humans or pets and are therefore not submitted often.

The submissions of *D. variabilis* and *A. americanum*, in contrast to *I. scapularis*, occurred almost exclusively during the summer. In June, 109 *D. variabilis* and 105 *A. americanum* were submitted, representing the peak submissions in both species. In July, 75 *D*. *variabilis* and 47 *A. americanum* were submitted; in August, 20 *D. variabilis* and 23 *A. americanum* were submitted. In contrast to *I. scapularis*, *A. americanum* is known to feed on a diverse array of mammals and birds at all three lifestages (35). Combined with its relatively large size, this makes it easier for humans to detect and submit at any lifestage.

Curiously, although *A. americanum* were submitted as both adults and nymphs, only adult *D. variabilis* were ever submitted. This suggests that while *A. americanum* is highly anthropophilic during multiple lifestages, *D. variabilis* is only anthropophilic during its adult stage with nymphs and larvae rarely being identified on humans or domestic animals. While adult *D. variabilis* are collected from a number of animals, the immature lifestages are more closely associated with small rodents (22,23). It is also possible that the larvae or nymphs were too small to detect, although this is unlikely considering the submission rate of *A. americanum* and *I. scapularis* larvae and nymphs which are of comparable size. *Ixodes cookei*, though rare, was detected with a similar temporal pattern as *D. variabilis* and *A. americanum*, with peak submissions occurring in June and throughout the summer.

The tick submissions changed substantially in the fall. October and November represent the peak of *I. scapularis* submissions, primarily as adults, with 871 submitted in October and 779 submitted in November. These ticks were nymphs during the summer that fed and molted to adults by mid-October. These ticks are also the same population that will overwinter and becomes prevalent in the subsequent spring. Therefore, the temporal distribution of adult *I. scapularis* is distinctly biphasic, with the highest risk of human exposure occurring in fall and spring. This consists of populations of adult ticks, which are the primary lifestage of *I. scapularis* submitted. Nymphs, which are more common during the summer, are submitted less often despite having a higher population in nature. This may be due to adults being more anthropophilic, while nymphs have a possibility of feeding on small rodents as larvae do. The size of nymphs may also make them more difficult to observe, although this is unlikely considering the size of an engorged nymph at the point of spontaneous detachment.

In contrast to *I. scapularis*, the number of *D. variabilis* and *A. americanum* submitted decreased drastically during the fall, with no *D. variabilis* submitted past August and only two *A. americanum* submitted in October. In New York, this period corresponds to slightly cooling temperatures, although frost is not typically present until November. This may be due to reduced human outdoor activity leading to a decreased occurrence of tick exposure, however this phenomenon is not observed for *I. scapularis*, which experiences peak submission numbers in October and November. It is more likely a factor of decreasing temperatures resulting in a decreased ability to quest or hunt, although the effect of changes to the day-night cycle have also been shown to influence tick questing behavior (36), with nocturnally questing *I. ricinus* increasing in activity as nights increase in length.

Tick submissions decreased as winter progressed, with only 52 *I. scapularis* being submitted in December. Tick activity during winter is unique to *I. scapularis*. These ticks are highly resistant to cold (37), but, unlike other species, are capable of actively questing during cold non-freezing conditions, hence their submission during December in the absence of all other species. Intermittent periods of temperatures above freezing are common during December in much of New York, and these data indicate that during these periods there is still a risk of tick exposure to *I. scapularis*.

Additionally, the repeated freezing and thawing of questing ticks during this period may increase tick mortality versus periods of continuous, consistent frost (38). This process related to energy storage and use by the tick rather than direct injury from freezing temperatures (39). Ticks have finite energy reserves, and using that energy during winter increases the risk of earlier spring mortality for ticks that fail to find a host.

### Tick Species Show Geographic Variation

The geographic distribution of ticks throughout the New York state is depicted by submissions per county in Figure 3. *Ixodes scapularis* is distributed consistently throughout the state, with the lowest numbers observed in the Southern Tier and Mohawk Valley/Adirondack regions. This may be due to the low population density of those areas. The greatest number of *I. scapularis* were submitted from the Hudson Valley/Catskill region and central New York (Albany, Columbia, Dutchess, Orange, and Ulster counties).

*D. variabilis* was primarily submitted from the Hudson Valley/Catskill region. This region is notable for thick forests but also a high human population, as well as more mild conditions than elsewhere in New York State. Many *D. variabilis* were also submitted from the western edge of the state (Niagara and Erie counties), and some from central New York in the vicinity of Syracuse (Onondaga and Cortland Counties). The actual distribution is most likely continuous, but this is not observable due to the low rate of submission in the more rural Northern and Finger Lakes regions.

*Amblyomma americanum* was identified throughout the state, with the primary center of its population being observed in Suffolk County on Long Island. As an island, this represents a geographically isolated environment; additionally, Suffolk County has a high population of both humans and deer, making it well suited both for the reproduction of ticks as well as their intersection with human life. *Amblyomma americanum* submissions from Suffolk County outnumbered *I. scapularis* submissions by 400%. This is indicative of a large and highly established tick population, although the additional encounters may be a result of differing behaviors between the ticks, such as greater anthropophilic tendencies in *A. americanum* or a tendency to inhabit different and more heavily populated areas of forests than *I. scapularis* normally does. There also appears to be a sizable distribution throughout mainland New York, with low numbers detected throughout the state’s southern and central portions. This pattern suggests that the tick is emergent or reemergent in the state and currently in the process of northward migration from its more traditional range south of the state. This agrees with published models indicating the possibility of New York state and the Northeast as a suitable habitat for *A. americanum* (40–42) and the detection of *A. americanum* in southern New England (43).

Although rare, the woodchuck tick, *I. cookei,* is of particular interest as a vector of Lineage I Powassan virus (44). It is uniquely elusive due to its close association to rodent burrows. However, submissions indicated that it is present in New York state sympatrically with its relative *I. scapularis* and that humans and domestic animals sometimes encounter it. Geographically, *I. cookei* were submitted from western and central New York, as well as the Adirondack region and southern Mohawk Valley. Some were additionally submitted from the Hudson Valley and Suffolk County, indicating that the species is present throughout the state.

*Haemaphysalis longicornis*, a species of tick endemic to Asia and recently introduced to the U.S., was not submitted in appreciable numbers. Therefore, gaining an understanding of its emergence into New York state and migration patterns is not possible from this dataset. Although it is present in New York state (45), it is rarely submitted; this may be due to rarity or due to host preference that does not include humans and pet species.

### Pathogen Distribution Corresponds to the Distribution of Tick Species

The majority of detected pathogens were those transmitted by *I. scapularis*. The most common pathogen detected in submitted ticks was *B. burgdorferi*. As shown in (Figure 2), *B. burgdorferi*-positive ticks were submitted from throughout the state. The rate was lowest in the Adirondack mountains and the Finger Lakes region. The number submitted was highest in the central New York and Hudson Valley/Catskill region. Temporally, the majority of *B. burgdorferi*-positive ticks were submitted in October (329 of 871 *I. scapularis*, 38%) and November (290 of 779 *I. scapularis*, 37%) (Figure 4). This suggests that the peak of *B. burgdorferi* positivity is in the fall, which corresponds with the emergence of adult ticks. These adults have had two chances to feed on potentially pathogen-positive rodent hosts during the summer (feeding as larvae and as nymphs), meaning they are the most likely lifestage to be infected with any pathogen.

**Figure 4.**
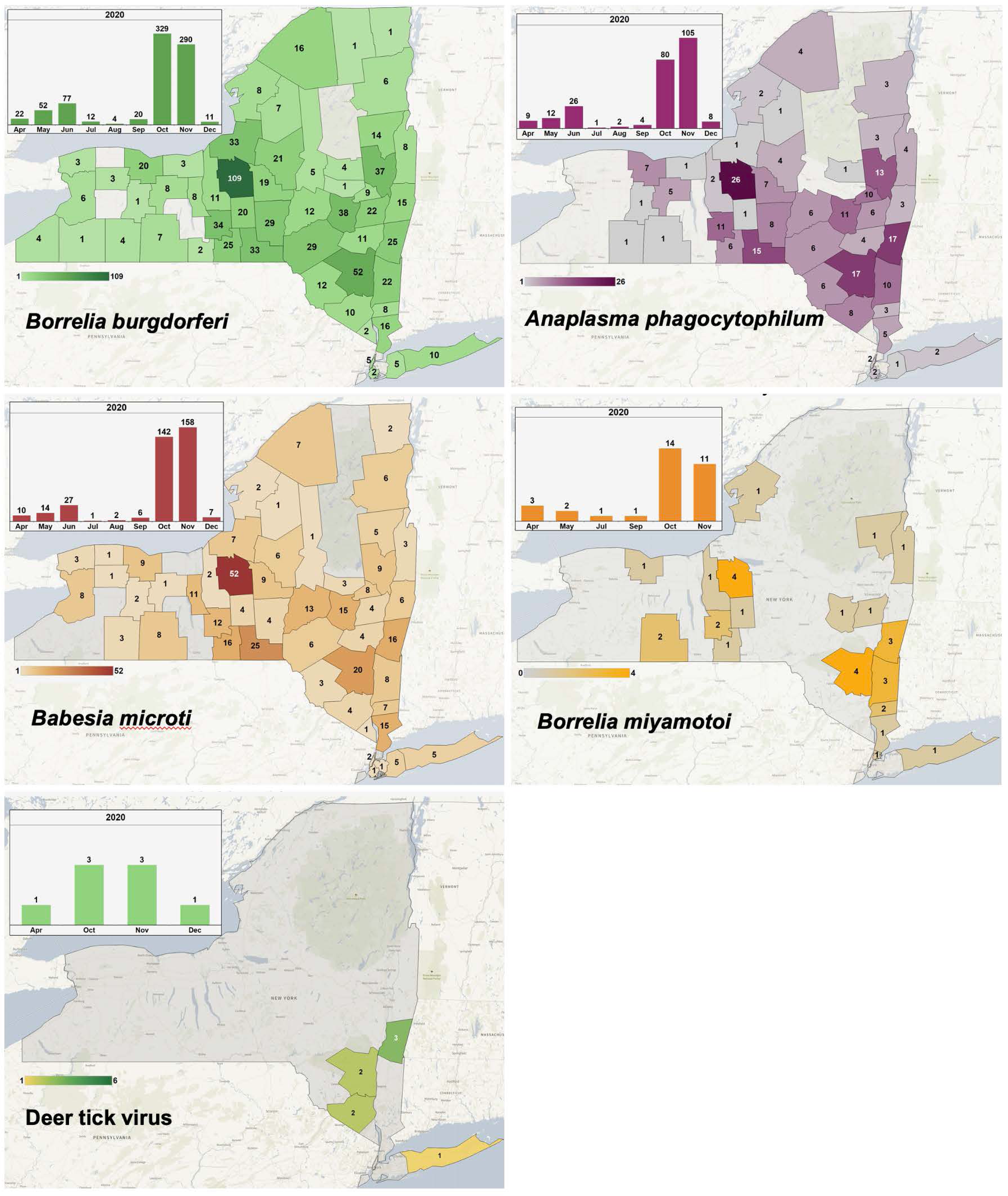
Prevalence of Tick Pathogens by County Level and the Seasonal Distribution of the Pathogens (inset). In New York, the most common pathogen in *I. scapularis* is *B. burgdorferi*, followed by *B. microti* and *A. phagocytophilum*, with statewide distribution. *Borrelia miyamotoi* is rarer but has a wide distribution across the state, while DTV, the least common pathogen observed, was restricted to the southern portion of the state and most prevalent in the Hudson Valley region.

Like *B. burgdorferi*, *A. phagocytophilum* was detected throughout the state, especially in the central and southern regions; also like *B. burgdorferi*, it’s period of highest positivity were observed in October (80/872 *I. scapularis*, 9%) and November (105/779 *I. scapularis*, 13%) (Figure 4). This trend is also mirrored in *B. microti*, with 142 (16% of *I. scapularis*) ticks observed in October and 158 (20% of *I. scapularis*) observed in November, as well as distributed evenly throughout the state. *Borrelia miyamotoi* shares a similar peak in autumn but is much rarer, with 14 ticks positive in October (2% of *I. scapularis*) and 11 in November (1% of *I. scapularis*). These ticks are also geographically related to the central New York area and the Hudson Valley, corresponding to regions with substantial numbers of submissions. The presence of these pathogens is directly related to the seasonal activity of *I. scapularis*.

POWV was rarely detected in submitted ticks. During the time-frame analyzed, all submitted ticks positive for POWV were identified as containing POWV Lineage II (Deer tick virus, DTV). These ticks were submitted from the more southern portion of the state in the Hudson Valley, with one from Suffolk County. One of these was observed in an overwintered adult in April, while the remainder were observed in adults from October, November, and December. This agrees with literature stating that adult *I. scapularis* are the primary vectors of DTV as opposed to nymphs (46,47,48), and further identifies that the period of greatest risk of exposure is fall.

Pathogens transmitted by non-*Ixodes* species were not observed in *D. variabilis* or *A. americanum*, suggesting that these pathogens are rarely encountered in New York and the probability of exposure is low. Pathogens associated with non-*Ixodes* species in this experiment include Heartland and Bourbon viruses as well as *Ehrlichia chaffeensis*. While Heartland virus is known to exist in New York (49), no human cases have been reported (50). Cases of Bourbon virus have additionally not been reported (51). *Ehrlichia chaffeensis*, transmitted by *A. americanum*, is present in New York at a rate of 7.81 cases per million (52), with all states in the Northeast reporting at least some cases and with an exceptionally high incidence of 62.5 cases per million in neighboring Vermont. This is a clear indication that the pathogen and the ticks that transmit it are present in New York but that encounters with them may be uncommon. This is most likely a result of the comparatively low number of *A. americanum* submitted, which makes the assessment of rare pathogens difficult. Additionally, it may be indicative of minimal human intersection with habitat conducive to maintaining the sylvatic cycle of *E. chaffeensis*. Thus, while *A. americanum* are present in New York, the subset containing *E. chaffeensis* may exist where human presence is infrequent.

### Coinfection was Detected at Various Rates with Up to Three Pathogens

Coinfections were observed in the submitted *I. scapularis* ticks. The majority of infected *I. scapularis* (40%) were infected with a single pathogen, although approximately 8% of submitted *I. scapularis* were infected with two pathogens and about 1% infected with three (Table 3). Fourth-order or higher coinfections were not observed. Rates of coinfection were predicted as the product of the rates of individual pathogens in singly-infected ticks.

**Table 3.** Coinfections Observed in *I. scapularis*. A list of the number of single-pathogen infections compared to the various combinations observed with two or three pathogens. No polymicrobial infections with greater than three pathogens were observed in this set of ticks. The majority of coinfections were associated with *B. burgdorferi*.

| Number of Pathogens | Pathogen Combination | Number of Ticks |
| --- | --- | --- |
| 1 | *B. burgdorferi* | 595 |
|  | *A. phagocytophilum* | 108 |
|  | *B. miyamotoi* | 15 |
|  | *Babesia microti* | 217 |
|  | Deer Tick Virus (DTV) | 4 |
| 2 | *A. phagocytophilum* / *B. miyamotoi* | 1 |
|  | *B. burgdorferi* / *A. phagocytophilum* | 89 |
|  | *B. burgdorferi* / *B. miyamotoi* | 6 |
|  | *B. burgdorferi* / *B. microti* | 101 |
|  | *B. microti* / *A. phagocytophilum* | 26 |
|  | *B. microti* / *B. miyamotoi* | 2 |
|  | *B. microti* / DTV | 1 |
| 3 | *B. burgdorferi* / *A. phagocytophilum* / *B. miyamotoi* | 5 |
|  | *B. burgdorferi* / *A. phagocytophilum* / DTV | 1 |
|  | *B. burgdorferi* / *B. miyamotoi* / DTV | 1 |
|  | *B. burgdorferi* / *B. microti* / *A. phagocytophilum* | 17 |
|  | *B. burgdorferi* / *Babesia microti* / *B. miyamotoi* | 2 |
|  | *B. burgdorferi* / *B. microti* / DTV | 1 |

The predicted and observed percentages of tick infection were generally similar, with some exceptions. For *A. phagocytophilum*, the predicted rate of single infection was 8.47%, although single infections were observed 3.81% of the time. This suggests that *A. phagocytophilum* occurs more often in ticks with another pathogen than it does alone.

A similar effect was observed for *B. microti*. Although the predicted rate of single infection was determined to be 12.4%, the observed rate of single-infection was lower at 7.66%. As with *A. phagocytophilum*, this suggests that *B. microti* in ticks occurs more often in coinfection with other pathogens than would be mathematically expected. 40.9% of all ticks with *B. microti* were infected with at least one other pathogen, with 33% of all *B. microti*-infected ticks being coinfected with *B. burgdorferi* as a constituent member of the co- or poly-infection.

DTV, although only detected in eight ticks within this timeframe, exhibited a high rate of coinfection. Four of the eight DTV-positive ticks were infected with another pathogen; in three of these, *B. burgdorferi* was a constituent component.

*Borrelia miyamotoi* was rarely detected, with a total rate of 1.13% in the *I. scapularis* population. Of these cases, 53.13% were infected with at least one additional pathogen. This indicates that *B. miyamotoi* commonly occurs in co- and poly-infections with other pathogens.

Several phenomena may be responsible for unexpected rates of coinfection. Firstly, they may be caused by the occurrence of direct interaction between the two pathogens, with inhibition or enhancement of one or both pathogens within the tick or changes to the success of pathogen acquisition or transmission. This has been identified with some pathogen pairs, with, for example, the presence of *A. phagocytophilum* altering the quantity of *B. burgdorferi* in mice to potentially enhance tick acquisition (53), or with *B. burgdorferi* coinfections enhancing acquisition, transmission, and ultimately assisting to sustain the sylvatic cycle of *B. microti* (54). This may result in specific pathogen pairs being overrepresented in ticks due to the symbiotic relationship of the two, or some pathogen pairs being rare due to competition for ticks and hosts.

The effect may also be due to variable sylvatic cycles sharing a tick vector but utilizing separate mammalian reservoirs. Although *P. leucopus* is understood to be a potential reservoir for a number of tick-borne pathogens (55), other less-common animals may be superior reservoirs for specific pathogens. These could belong to a large array of birds, reptiles, and mammals, including many small rodents such as jumping mice (*Napaeozapus insignis*), voles (*Microtus* spp.), and sciurids (*Tamias striatus*, *Sciurus carolinensis*) that may serve an analogous role to *P. leucopus* as a tick host in different settings. Pathogen enhancement due to coinfection, therefore, may be associated either with overlapping ranges of specific hosts or with hosts that support multiple pathogens. Less common coinfection, likewise, may be the result of two reservoirs that each only support the growth of one pathogen failing to share a habitat.

Consideration of coinfections in ticks and their likeliness is important to developing treatments for clinically observed co-infections of tick-borne diseases. Treatment for Lyme disease, for example, uses doxycycline, amoxicillin, or cefuroxime (56). Of these antibiotics, only doxycycline is considered highly effective against *A. phagocytophilum* and amoxicillin is considered mostly ineffective (57). Additionally, as a eukaryote, *B. microti* requires specific treatment using azithromycin or clindamycin with quinine (58), requiring more complex treatment when paired with bacteria. Infections involving multiple pathogens may also result in unique disease states with variable clinical presentation.

The prevalence of coinfections involving rare pathogens (DTV and *B. miyamotoi*) are more difficult to analyze due to the low rate of occurrence of these pathogens and consequently low detection of coinfection per year.

### Limitations of the Community-Science Submission Program

Community-surveillance programs have often been used to track the emergence and persistence of tick populations and tick-borne pathogens (59–68). These vary in scale and methodology. In some cases, the ticks themselves are not collected, with information being instead gained from photographs of ticks submitted for identification (60,65). This process, although requiring less reagent and processing investment, relies heavily on the quality of the photographs taken especially with regard to fine-detail necessary for species identification within a genus (69) but can be more accessible to users unwilling to handle or mail tick samples. Other app-based systems allow for submission of data concerning tick contact only, which can be used to deliver educational materials and to map with tick/human contact (67,66).

Other programs exist that allow tick submission for identification and pathogen testing. In most cases, these are intended to focus on a specific set of medically important pathogens associated with a geographic area, although the system utilized can be quickly retooled to handle different pathogens and ticks in different locations (68). In North America, these often include *B. burgdorferi*, *A. phagocytophilum*, *B. microti*, and *B. miyamotoi* (59,68) but in some cases are restricted to *Borrelia* species only (70,71). For European programs, where *I. ricinus* are the predominant tick species, the pathogens of interest vary; in a Belgian study, the targets included *Babesia spp.*, *A. phagocytophilum*, *B. miyamotoi*, *Neorickettsia mikurensis,* and *Rickettsia Helvetica*, with *D. reticulatus* ticks being tested for *R. raoultii* (72). Tests were also conducted for tick-borne encephalitis virus (TBEV). A different study concerning ticks submitted specifically by wild boar hunters in southern Italy focused on more variable tick species, including *D. marginatus*, *Rhipicepualus sanguineus*, *R. turanicus*, and only a small number of *I. ricinus*, with the pathogen targets being primarily *Rickettsia* species in addition to *Coxciella burnetiid*, *B. lusitaniae*, and *Candidatus* Midichloria mitichondrii (73).

An important limitation in programs that screen for pathogens is the use of DNA-based identification, either through PCR or by sequencing. While this allows for testing of all tick-borne prokaryotic and eukaryotic pathogens, it ignores the presence of tick-borne viruses which primarily utilize RNA genomes and produce intermediate DNA during replication. In these cases, the RNA must be extracted with a second step, as for European programs that test for TBEV (72). This results in greater procedural complexity and reagent cost for virus testing, and as such, viruses are usually ignored.

Community-science initiatives have also been applied toward the development of maps. These can be used to map pathogen occurrence (59,61) or can also be utilized without pathogen data to determine the distribution of tick species (62,63,64). These maps can be highly detailed and contain the summaries of substantial datasets (64) although are often limited by finding a means to efficiency disseminate the mapping information. This limits the ability for the public to respond to risks assessed and mapped by the investigation

There are several fundamental limitations to the type of community-driven submission-based approach used for this study. These include limitations concerning accuracy of data acquisition and internal biases derived from the method allowed for tick collection, although many of these limitations are reduced for programs utilizing submitted samples versus photographs (69). Firstly, in this system, the first portion of the data acquired (location, date, host origin, and biting state) is received through a survey presented to the tick submitter. Consequently, this data is provided directly by the public. This is less controlled than the tick identification and pathogen screening, which occur in a laboratory setting, and is subject to unverifiable error. This most commonly would include marking a tick found crawling on the body as having been attached when it has not yet bitten or providing potentially inaccurate information about where or when the tick was found. This especially introduces uncertainty when a tick is found after a person has been travelling, resulting in an imprecise or inaccurate representation of the tick’s true origin.

Additionally, the submitter is responsible for packaging and sending the tick and as such the ticks can arrive in various states of integrity. The submission form recommends submitting live ticks in sealed bags with moist paper to ensure survival, although ticks may also be submitted dead in alcohol or formalin and sometimes arrive dead, fully desiccated, missing pieces, or moldering. The tick condition is recorded upon receipt, although damaged ticks can reduce the possibility of accurate species identification and can decrease the quality of material for qPCR pathogen detection and future culturing efforts.

This is especially important for an RNA-based assay. Although this process allows for straightforward detection of viral pathogens in addition to prokaryotic and eukaryotic ones, it relies on RNA, which is less stable than DNA over time. Severely degraded RNA may result in false-negative readings. Attempts have been made to combat this issue by providing instructions to potential submitters that guide packaging the ticks to ensure the best possible results and by the planned addition of a tick-RNA based primer set in the qPCR multiplex panel to assess the quality of the RNA during screening.

The information produced by this type of community-driven process is also restricted in that it concerns only populations of ticks that come in contact with humans or human-associated animals. This is partially advantageous in that it relates to actual human risk more directly than active surveillance. This is especially true in cases where a pathogen may be present in the state but restricted to specific sylvatic cycles, such as undisturbed woodland environments, where few humans go. This may result in some pathogens, such as *E. chaffeensis* or POWV/DTV, being detected infrequently or not at all despite the known existence of these pathogens in the state. The disadvantage of a community-based submission approach, however, is that the ticks sampling is widely dispersed over a number of habitats and locations distributed throughout the state. This means that unless many ticks are submitted from a small area, the data fails to capture variability in tick and pathogen density based on landscape features. An area with a forested park, for example, may have a high pathogen prevalence within the park but not in its suburban areas; these rates, however, would be averaged over a zip-code or county sized region and thereby failing to account for these features.

Furthermore, human contact with ticks is directly related to human outdoor activity. Assessing tick activity, therefore, is not possible from this data, as human outdoor activity and submissions are not consistent throughout the year. Summer, for example, has a different level of outdoor activity than winter, and the survey questionnaire does not currently address this. Likewise, the number of ticks submitted is directly proportional to public enthusiasm. This may result in year-by-year variation in the number of ticks submitted as the program gets more or less popular, or sudden smaller increases following news coverage or other advertisement of the program.

Despite these limitations, the community submission approach to surveillance has resulted in a set of data derived from the entirety of New York state throughout 2020, in a program that can continue over multiple years for additional temporal data from the state. This provided data on the tick and tick-borne pathogen presence throughout the state on a scale that would not be feasible using active surveillance. This data, in turn, was used to develop a mapping tool available to residents throughout New York to assist in determining their risk of encountering specific species of ticks or varieties of tick-borne pathogens.

## CONCLUSION

A Citizen Science community engagement program was utilized to acquire ticks from throughout New York state in 2020. These ticks were categorized by species and screened for a panel of several tick-borne pathogens. This data was then combined with their origin, as identified by the tick submitter on the submission survey, to create a database of ticks, their species, their pathogen status, and other data concerning them throughout the state. The primary species observed included *I. scapularis*, *D. variabilis*, and *A. americanum*, the latter of which is in the process of colonizing the southern portions of the state. Low levels of *I. cookei* and non-*scapularis Ixodes* were also observed. Of the observed species, only *I. scapularis* was commonly infected with pathogens of human relevance, including *B*. *burgdorferi*, *B. miyamotoi*, *A. phagocytophilum*, *B. microti*, and POWV/DTV. The host origin of submitted ticks was also found to be mostly for humans and domestic animals that humans commonly encounter, including cats and dogs.

This project has allowed for the construction of a powerful interactive mapping dashboard, *tick*MAP, that allows for community engagement, including allowing individuals to assess and track the presence of ticks and the risk of tick-borne diseases to a county level. This same data may also be used as a practical or educational resource for clinicians of both human and animal patients for developing differential diagnosis and informing clinical care. It has also identified several areas of high-pathogen density for more intensive field surveillance and for acquiring ticks of unusual and under-studied species for laboratory colonization. Over time, it will allow for the identification of changes to tick-borne pathogen distribution as well as the distribution and emergence of tick species themselves. This can in turn be related to other factors, such as human population, yearly climatic variation, long-term climate alteration, forest type, animal behavior, and land development patterns to gain a better understanding of the cause of tick expansion and the role of extrinsic factors in the transmission and cycling of tick-borne pathogens. In conclusion, we have developed a powerful health tool to track the emergence of ticks and tick-borne disease-causing agents. This public health tool could be adapted to track the emergence of ticks and tick-borne agents for other states or territories or countries.

## Data Availability

All data produced in the present work are contained in the manuscript

http://www.nyticks.org

http://www.tickmap.org

## ACKNOWLEDGMENTS

We would like to acknowledge the collaborations of our IMT Research Support staff and Moonshot team in developing a workflow and technical process for data gathering and the Tableau dashboards. We also would like to thank Ms. Alica Quattropani for her help with tick identifications.

## DISCLAIMERS

The funders do not have any role in the experimental design and interpretation of the results described in this manuscript.

## AUTHOR BIO

Dr. Hart is a postdoctoral fellow in the Department of Microbiology and Immunology at SUNY Upstate Medical University. His research interest focuses on the transmission and biology of arthropod-borne pathogens, particularly in the context tick- and mosquito-borne viruses and coinfection with multiple pathogenic agents.

---

**Supplementary Figure 1.**
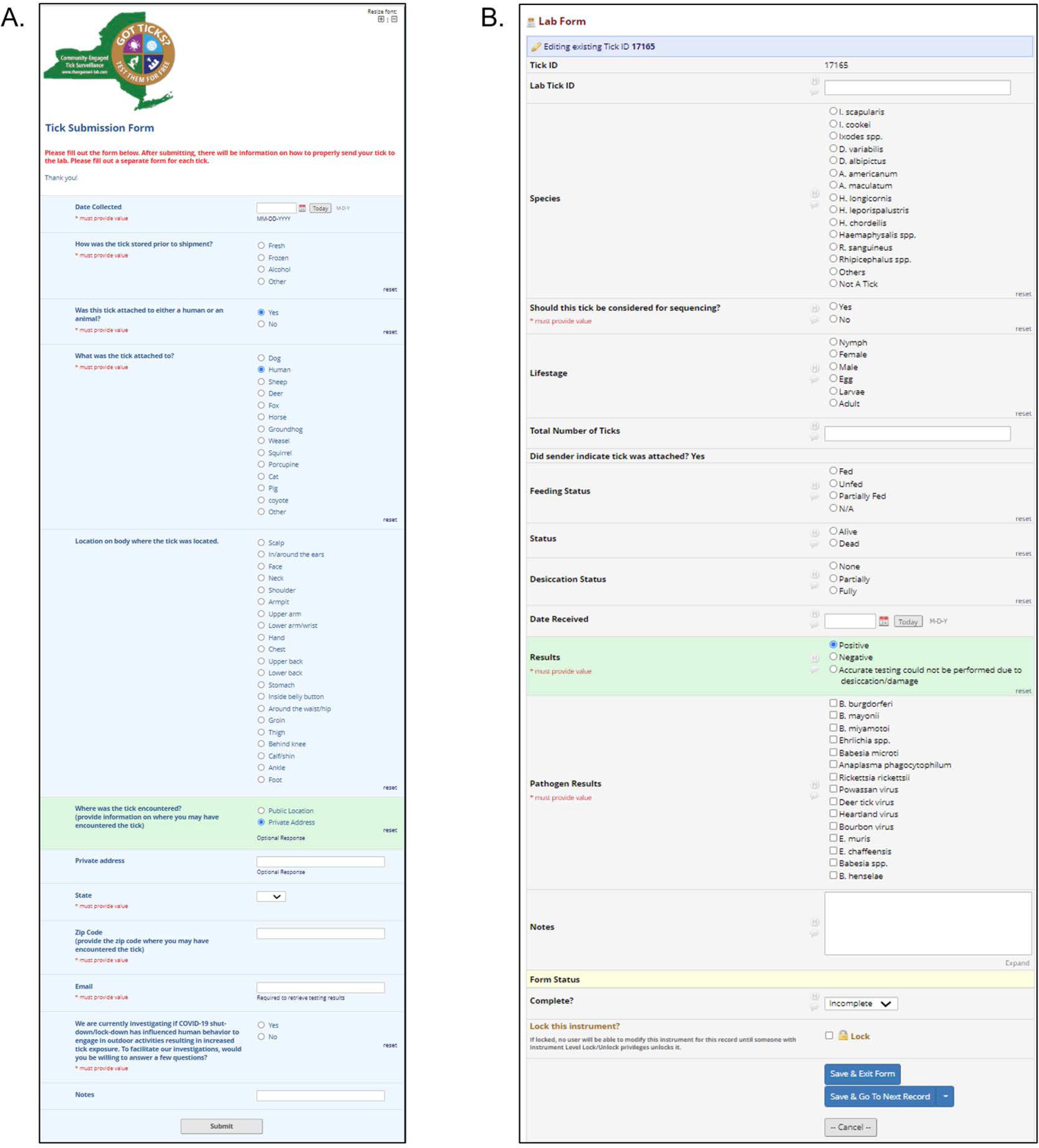
Data collection and results forms. Two-part survey in REDCap. Tick Submission Form (A), completed by tick submitters, and the Lab Form (B), completed by technical staff.

